# SARS-CoV-2 anti-spike antibody levels following second dose of ChAdOx1 nCov-19 or BNT162b2 in residents of long-term care facilities in England (VIVALDI)

**DOI:** 10.1101/2022.01.26.22269885

**Authors:** Oliver Stirrup, Maria Krutikov, Gokhan Tut, Tom Palmer, David Bone, Rachel Bruton, Chris Fuller, Borscha Azmi, Tara Lancaster, Panagiota Sylla, Nayandeep Kaur, Eliska Spalkova, Christopher Bentley, Umayr Amin, Azar Jadir, Samuel Hulme, Rebecca Giddings, Hadjer Nacer-Laidi, Verity Baynton, Aidan Irwin-Singer, Andrew Hayward, Paul Moss, Andrew Copas, Laura Shallcross

## Abstract

**Background:** General population studies have shown strong humoral response following SARS-CoV-2 vaccination with subsequent waning of anti-spike antibody levels. Vaccine-induced immune responses are often attenuated in frail and older populations such as Long-Term Care Facility (LTCF) residents but published data are scarce.

**Methods:** VIVALDI is a prospective cohort study in England which links serial blood sampling in LTCF staff and residents to routine healthcare records. We measured quantitative titres of SARS-CoV-2 anti-spike antibodies in residents and staff following second vaccination dose with ChAdOx1 nCov-19 (Oxford-AstraZeneca) or BNT162b2 (Pfizer-BioNTech). We investigated differences in peak antibody levels and rates of decline using linear mixed effects models.

**Results:** We report on 1317 samples from 402 residents (median age 86 years, IQR 78-91) and 632 staff (50 years, 37-58), ≤280 days from second vaccination dose. Peak antibody titres were 7.9-fold higher after Pfizer-BioNTech vaccine compared to Oxford-AstraZeneca (95%CI 3.6-17.0; *P*<0.01) but rate of decline was increased, and titres were similar at 6 months. Prior infection was associated with higher peak antibody levels in both Pfizer-BioNTech (2.8-fold, 1.9-4.1; *P*<0.01) and Oxford-AstraZeneca (4.8-fold, 3.2-7.1; *P*<0.01) recipients and slower rates of antibody decline. Increasing age was associated with a modest reduction in peak antibody levels for Oxford-AstraZeneca recipients.

**Conclusions:** Double-dose vaccination elicits robust and stable antibody responses in older LTCF residents, suggesting comparable levels of vaccine-induced immunity to that in the general population. Antibody levels are higher after Pfizer-BioNTech vaccination but fall more rapidly compared to Oxford-AstraZeneca recipients and are enhanced by prior infection in both groups.

## Introduction

Residents of Long-Term Care Facilities (LTCF) have experienced extremely high rates of SARS-CoV-2 infection and mortality[1]. Since December 2020, LTCF staff and residents in England have been prioritised for vaccination against SARS-CoV-2, with initial roll-out primarily using the mRNA-based BNT162b2 (Pfizer-BioNTech) and adenoviral vector-based ChAdOx1 (Oxford-AstraZeneca) vaccines[2].

Vaccine effectiveness in the general population has been demonstrated for at least six months following second dose administration[3,4]. However, data are limited on the duration and magnitude of protection afforded by vaccination in LTCF residents. Furthermore, LTCF residents are especially vulnerable to severe outcomes following infection due to frailty, high rates of co-morbidity, poorer nutritional status, and age-related dampening of immune responses (immune-senescence) which impact on vaccine-induced immunity[5,6].

Current SARS-CoV-2 vaccines target the viral spike protein, and anti-spike antibody levels are an important correlate of vaccine efficacy[7]. Early studies are encouraging and suggest robust cellular and humoral responses in the initial months following vaccination amongst LTCF residents, particularly in previously-infected individuals[8–10]. However, studies from the general population have reported waning of antibody titres in the six months following vaccination, particularly in people older than 65 years[11–13]. We investigated quantitative anti-spike antibody titres amongst LTCF staff and residents in England over the first nine months following second vaccination dose.

## Methods

VIVALDI (ISRCTN 14447421) is a prospective cohort study of residents and staff of LTCFs in England[14]. Eligible individuals from participating LTCFs provide written informed consent for study participation and consultees are sought for residents lacking capacity to consent. Participants have undergone up to five rounds of blood sampling at eight-week intervals between 11 June 2020 and 22 October 2021. As part of the national pandemic response, all LTCF staff and residents regularly submit nasopharyngeal swabs for SARS-CoV-2 PCR testing (monthly in residents, weekly in staff) with additional testing during outbreaks[15]. Blood samples undergo SARS-CoV-2 nucleocapsid IgG testing using the Abbott ARCHITECT semi-quantitative immunoassay (Maidenhead, UK). Quantitative antibody titres against SARS-CoV-2 spike and nucleocapsid IgG are measured using the Meso Scale Diagnostics (MSD) V-PLEX COVID-19 Respiratory Panel 2 kit (Rockville, MD, USA). Anti-nucleocapsid antibodies are used to identify immune responses stimulated by prior infection[16]. MSD observations were included from ≥21 days after second vaccine dose administration, corresponding to peak antibody response[17], up until date of third vaccine dose where recorded. Only individuals with data on demographic characteristics and vaccinations were included in this analysis and most could also be linked to full testing history (Appendix S1).

To model post-vaccination MSD assay anti-spike antibody levels, individuals were categorised as either having ‘no evidence of prior infection’ or ‘evidence of prior infection’. The latter group included individuals with at least one record of an active infection defined by PCR or point-of-care lateral flow test (LFT) positivity or hospitalisation with COVID-19 prior to second vaccine dose, and those with presence of anti-nucleocapsid antibodies on either Abbott or MSD assay. To exclude breakthrough infections which may have boosted antibody levels, observations with active infection recorded after second vaccine dose but prior to index date were dropped from analysis, as were observations following post-vaccination anti-nucleocapsid seroconversion.

An index value ≥0.8 defined Abbott anti-nucleocapsid assay positivity[18,19]. A threshold of 1200 AU/mL was used for MSD anti-nucleocapsid assay, which had a specificity of 96% (48/50) using pre-pandemic blood samples.

VIVALDI has been granted research ethics approval by the South Central-Hampshire B Research Ethics Committee (ref:20/SC/0238).

### Statistical analysis

Log10-transformed MSD anti-spike levels were modelled using linear mixed effects models. Time was centred at 21 days after second vaccine dose, with random intercept and slope terms for each participant.

An initial model was fitted with independent effects assumed for vaccine type, sex, staff/resident status and prior SARS-CoV-2 infection, followed by a model with interaction terms between vaccine type and each other variable. A further model was considered with addition of subject-age (centred at 70 years) as a linear predictor of both intercept and slope by vaccine type. Half-life values were calculated based on estimated time to drop in mean log10 antibody level of log10(0.5). Formal sample size calculation was not undertaken.

## Results

We describe 558 anti-spike antibody (MSD) results from 402 LTCF residents and 759 from 632 staff. 774 people had one observation, 237 had two and 23 had three. Median age was 86 (IQR 78-91) years for residents and 50 (IQR 37-58) years for staff. Median time from second vaccine dose to blood sample was 136 days (IQR 104-170, range 21-280). Four observations from four residents and four from three staff were dropped from analysis as they followed breakthrough infection. Eight residents and eight staff each had one observation excluded because of indirect evidence of breakthrough infection (i.e., appearance of anti-nucleocapsid antibodies).

The interaction model, allowing different effects by vaccine type, was found to provide better fit to the data than the simpler independent effects model (P=0.01, likelihood ratio test (LRT)), and a further improvement was found by adding age as linear predictor of peak antibody levels and slope (P=0.03, LRT).

Peak antibody titres were greater in Pfizer-B recipients than in Oxford-AZ recipients (×7.9, 95%CI 3.6-17.0; *P*<0.01), although we also observed a steeper annual decline in this group (×0.08 at 12 months *vs* equivalent decline from peak, 0.01-0.72; *P*=0.02) (Table 1, Figure 1). Prior infection with SARS-CoV-2 was associated with higher peak antibody levels and slower decline for both Pfizer-B (peak ×2.8, 1.9-4.1; *P*<0.01) and Oxford-AZ (×4.8, 95%CI 3.2-7.1; *P*<0.01) recipients. Male sex was associated with slightly higher peak in antibody levels for both vaccines (not statistically significant) but steeper decline, particularly for Oxford-AZ recipients. LTCF resident vs staff status was not associated with any statistically significant difference in peak antibody level or slope of decline. However, increasing age was associated with lower antibody peak for Oxford-AZ recipients.

**Table 1.** Regression coefficients from final statistical model for anti-spike antibody levels from 21 days following second vaccine dose, fitted to log10-transformed data

|  | *n, n (%)* or median (IQR) | *Intercept<sup>†</sup> (95%CI); P* | *Slope (95% CI); P [annual change]* |
| --- | --- | --- | --- |
| Reference coefficients‡ |  | 4.12 (3.86 to 4.38) | -0.67 (-1.48 to 0.14) |
| *Oxford-AZ recipients* | 493 | *Difference in intercept (95%CI)#; P* | *Difference in slope (95%CI)\*\*; P* |
| Prior infection (yes vs no) | 246 (49.9) | 0.68 (0.5 to 0.85); <0.01 | 0.50 (-0.01 to 1.01); 0.06 |
| LTCF resident (vs staff) | 251 (50.9) | 0.22 (-0.14 to 0.59); 0.23 | -0.45 (-1.58 to 0.67); 0.43 |
| Male (vs female) | 105 (21.3) | 0.17 (-0.05 to 0.39); 0.13 | -0.69 (-1.32 to -0.05); 0.03 |
| Age (per 10y greater than 70) | 67 (48–87) | -0.10 (-0.18 to -0.02); 0.01 | 0.16 (-0.09 to 0.42); 0.20 |
| *Pfizer-B. recipients* | 534 | *Difference in intercept (95%CI)#; P* | *Difference in slope (95%CI)\*\*; P* |
| Difference vs Oxford-AZ¶ |  | 0.90 (0.56 to 1.23); <0.01 | -1.09 (-2.04 to -0.14); 0.02 |
| Prior infection (yes vs no) | 306 (57.3) | 0.44 (0.27 to 0.61); <0.01 | 0.43 (0.01 to 0.85); 0.04 |
| LTCF resident (vs staff) | 147 (27.5) | -0.05 (-0.36 to 0.26); 0.74 | 0.06 (-0.7 to 0.82); 0.87 |
| Male (vs female) | 94 (17.6) | 0.11 (-0.1 to 0.31); 0.31 | -0.23 (-0.72 to 0.26); 0.36 |
| Age (per 10y greater than 70) | 56 (44–71) | -0.01 (-0.08 to 0.06); 0.76 | -0.06 (-0.23 to 0.11); 0.49 |

LTCF, long-term care facility.
\*% calculated using number with same vaccine type as denominator.
†Representing average peak value at 21 days after second vaccine dose.
‡Values for Oxford-AZ recipient female staff member at 70 years of age without prior infection. ¶Taken alone, represents the difference for female staff member at 70 years of age without prior infection.
#10<sup>x</sup> gives multiplicative difference in intercept associated with each factor.

**Figure 1:**
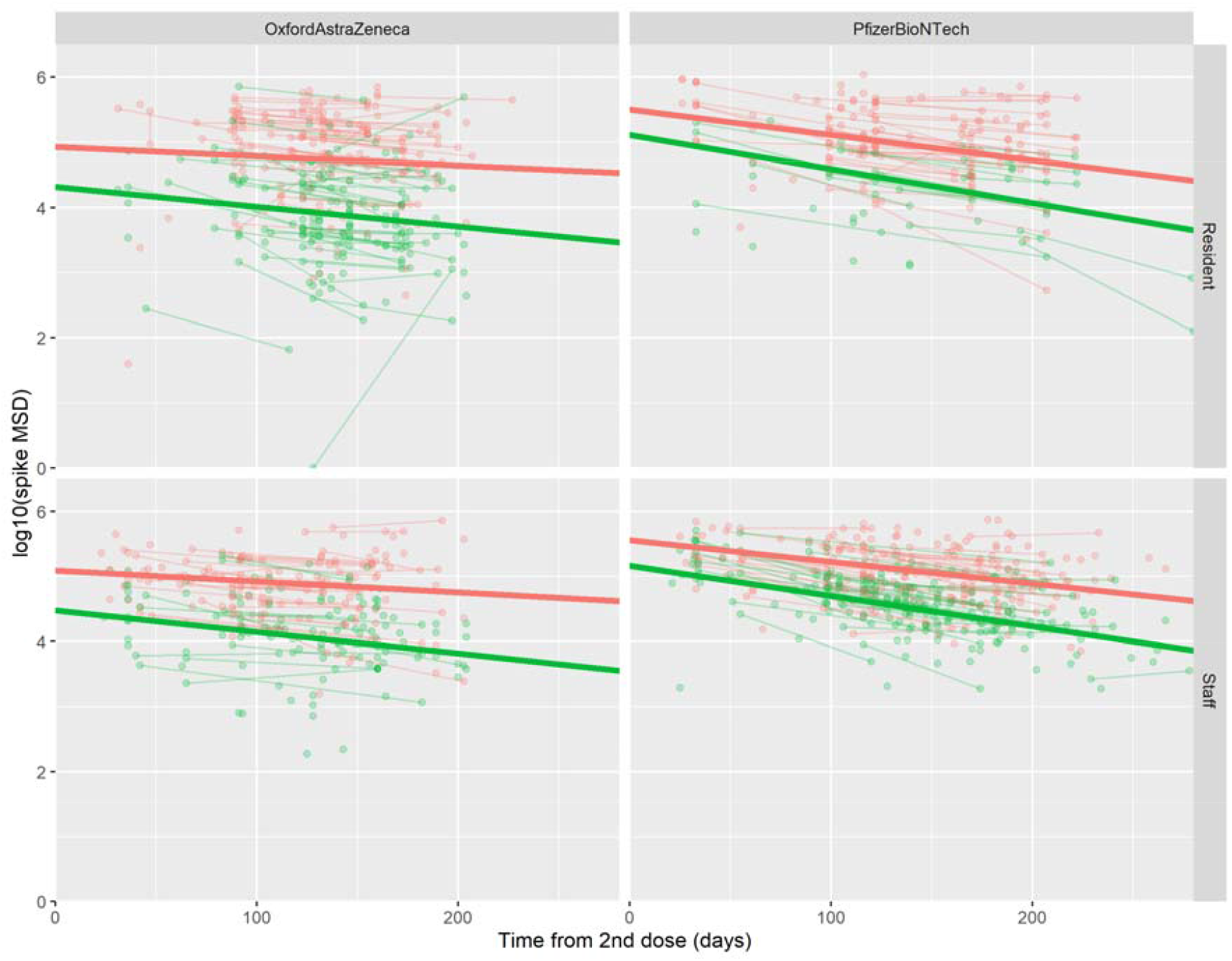
Log-transformed MSD values for anti-spike antibody levels in relation to the time from second vaccine dose, divided by vaccine type and staff/resident status, and colour-coded by prior infection category (red: evidence of prior infection; green: no evidence of prior infection). Individual observations are shown as dots, with those from the same person linked by lines. The bold straight lines show regression fits from a statistical model (omitting age and sex) to estimate trends in each group.

‘Half-life’ estimates of antibody decline were in the range 60-120 days for most subgroups, with values >6 months in female Oxford-AZ recipients with prior infection, but 95% CIs were wide (Table S2).

## Discussion

We present post-vaccination serological data from a large cohort of frail LTCF residents in England, a group in whom published data are scarce. Our findings are broadly consistent with longitudinal studies conducted in the general population and healthcare workers[11,13] which is reassuring given the vulnerability of LTCF residents to SARS-CoV-2 infection.

Consistent with previous studies, we find higher peak antibody titres following vaccination with Pfizer compared to Oxford-AZ[12,13]. Wei et al reported on anti-spike antibody waning in ∼100,000 Oxford-AZ and ∼55,000 Pfizer-B vaccine recipients, sampled through the Coronavirus Infection Survey (CIS)[13,20]. For Oxford-AZ they found peak antibody levels were higher in those with prior infection, and slightly lower in males and younger ages. Peak antibody levels were greater in Pfizer-B recipients compared with Oxford-AZ but were lower at older ages and for males[13].

The collection of samples up to 9 months after vaccination allowed us to assess the rate of spike-specific antibody decline from peak value. The mean half-life of antibody decline was reported as 85 days (95%CI 84-86) after Oxford-AZ in the CIS study, and this was increased to 131 days in those with prior infection. They found comparable mean half-life after Pfizer-B of 101 days (100-102) which was extended to 188 days in those with prior infection[13]. Our data also revealed mean half-life in the range 60-120 days but did not uncover significant variation in the rate of antibody decline between LTCF staff and residents. Analysis of >8500 community-dwelling infection-naïve adults also found no difference in rates of waning in donors aged ≥65 years although peak titres declined with age[12].

Our study is consistent in finding higher peak levels and longer half-life associated with prior infection for both vaccine types, and higher peak levels following Pfizer vaccination. Overall, our results are encouraging and add to a body of evidence suggesting strong humoral and cellular responses to vaccination amongst LTCF residents[9,10].

Our study is limited by a modest sample size, so there is uncertainty regarding the presence and magnitude of observed effects. It is also possible that some individuals labelled as infection-naïve may have waned below the positivity threshold following infection early in the pandemic[21]. To account for this, we used a lower Abbott positivity threshold and included MSD results in defining “prior-exposure”, but we cannot determine the chronology of infection in anti-nucleocapsid antibody positive participants. Finally, we have only described humoral responses to vaccination; analyses of vaccine-induced cellular immune responses in LTCF staff and residents are underway by our group and others.

Insights into the magnitude and duration of vaccine-induced immune responses are crucial to inform the timing of booster vaccination, particularly with the emergence of novel variants such as Omicron. Our findings reveal that current COVID-19 vaccines retain high immunogenicity in the LTCF setting but factors such as peak antibody response and rate of antibody waning, which will be used to guide the need for future vaccinations, are strongly influenced by vaccine regimen and prior infection status. Ongoing assessment of humoral immunity will be important in order to guide introduction of optimal booster regimens that maintain immunity over the longer term.

## Data Availability

De-identified test results and limited metadata will be made available for use by researchers in future studies, subject to appropriate research ethical approvals once the VIVALDI study cohort has been finalised. These datasets will be accessible via the Health Data Research UK Gateway.

## Acknowledgements

We thank the staff and residents in the LTCFs that participated in this study and Mark Marshall at NHS England who pseudonymised the electronic health records.

## References

1. ECDC. Surveillance data from public online national reports on COVID-19 in long-term care facilities. 2021.

2. http://GOV.UK. Vaccine update: issue 315, December 2020, COVID-19 special edition - http://GOV.UK.

3. Andrews N, Tessier E, Stowe J et al. Vaccine effectiveness and duration of protection of Comirnaty, Vaxzevria and Spikevax against mild and severe COVID-19 in the UK. medRxiv 2021:2021.09.15.21263583.

4. Thomas SJ, Moreira ED, Kitchin N et al. Safety and Efficacy of the BNT162b2 mRNA Covid- 19 Vaccine through 6 Months. New England Journal of Medicine 2021;385:1761–73.

5. Gordon AL, Franklin M, Bradshaw L et al. Health status of UK care home residents: a cohort study. Age and ageing 2014;43:97–103.

6. Castro-Herrera VM, Lown M, Fisk HL et al. Relationships Between Age, Frailty, Length of Care Home Residence and Biomarkers of Immunity and Inflammation in Older Care Home Residents in the United Kingdom. Frontiers in Aging 2021;0:2.

7. Feng S, Phillips DJ, White T et al. Correlates of protection against symptomatic and asymptomatic SARS-CoV-2 infection. Nature Medicine 2021 27:11 2021;27:2032–40.

8. Parry H, Bruton R, Tut G et al. Immunogenicity of single vaccination with BNT162b2 or ChAdOx1 nCoV-19 at 5–6 weeks post vaccine in participants aged 80 years or older: an exploratory analysis. The Lancet Healthy Longevity 2021;2:e554–60.

9. Tut G, Lancaster T, Butler MS et al. Reduced Antibody and Cellular Immune Responses Following Dual COVID-19 Vaccination Within Infection-Naïve Residents of Long-Term Care Facilities. SSRN Electronic Journal 2021, DOI: 10.2139/SSRN.3979590.

10. Tut G, Lancaster T, Krutikov M et al. Profile of humoral and cellular immune responses to single doses of BNT162b2 or ChAdOx1 nCoV-19 vaccines in residents and staff within residential care homes (VIVALDI): an observational study. The Lancet Healthy Longevity 2021;2:e544–53.

11. Levin EG, Lustig Y, Cohen C et al. Waning Immune Humoral Response to BNT162b2 Covid-19 Vaccine over 6 Months. New England Journal of Medicine 2021;385:e84.

12. Aldridge RW, Yavlinsky A, Nguyen V et al. Waning of SARS-CoV-2 antibodies targeting the Spike protein in individuals post second dose of ChAdOx1 and BNT162b2 COVID-19 vaccines and risk of breakthrough infections: analysis of the Virus Watch community cohort. medRxiv 2021:2021.11.05.21265968.

13. Wei J, Pouwels KB, Stoesser N et al. SARS-CoV-2 anti-spike IgG antibody responses after second dose of ChAdOx1 or BNT162b2 and correlates of protection in the UK general population. medRxiv 2021:2021.09.13.21263487.

14. Krutikov M, Palmer T, Donaldson A et al. Study Protocol: Understanding SARS-Cov-2 infection, immunity and its duration in care home residents and staff in England (VIVALDI). Wellcome Open Research 2020;5:232.

15. http://GOV.UK. Coronavirus (COVID-19) testing in adult care homes. 2021.

16. Houlihan CF, Beale R. The complexities of SARS-CoV-2 serology. The Lancet Infectious Diseases 2020;20:1350–1.

17. Eyre DW, Lumley SF, Wei J et al. Quantitative SARS-CoV-2 anti-spike responses to Pfizer–BioNTech and Oxford–AstraZeneca vaccines by previous infection status. Clinical Microbiology and Infection 2021;27:1516.e7–1516.e14.

18. Bryan A, Pepper G, Wener M et al. Performance Characteristics of the Abbott Architect SARS-CoV-2 IgG Assay and Seroprevalence in Boise, Idaho. Journal of clinical microbiology 2020;58, DOI: 10.1128/JCM.00941-20.

19. Ainsworth M, Andersson M, Auckland K et al. Performance characteristics of five immunoassays for SARS-CoV-2: a head-to-head benchmark comparison. The Lancet Infectious Diseases 2020;20:1390–400.

20. ONS. Coronavirus (COVID-19) Infection Survey, UK Statistical bulletins.

21. Krutikov M, Palmer T, Tut G et al. Prevalence and duration of detectable SARS-CoV-2 nucleocapsid antibodies in staff and residents of long-term care facilities over the first year of the pandemic (VIVALDI study): prospective cohort study in England. The Lancet Healthy Longevity 2021;0:2021.

