## Supplementary appendix for "SARS-CoV-2 anti-spike antibody levels following second dose of ChAdOx1 nCov-19 or BNT162b2 in residents of long-term care facilities in England (VIVALDI)"

Appendix S1

[**Figure S6:** Abbott anti-nucleocapsid antibody titres vs time since most recent known active infection (horizontal lines=log10(0.8) and log10(1.4)) [104 observations from 78 residents and 45 from 34 staff]	11](#_Toc94072790)

### **Data linkage**

Individuals are allocated a pseudo-identifier based on their National Health Service (NHS) unique identifier. Data linkage using the pseudo-identifier is undertaken in the NHS COVID-19 Datastore, established during the pandemic by NHS England for secure data storage (link). Results of PCR testing are linked to dates of vaccination from the National Immunisation Management System (NIMS), and hospital admission codes and dates from the Hospital Episode Statistics (HES) dataset. Blood sample results are processed by NHS England and linked by pseudo-identifier. Additional vaccination data was obtained directly from the care providers in cases where data linkage was unsuccessful.

**Case classifications**

We have divided post-vaccination MSD observations into the following categories for reporting and data visualisation:

1. No evidence past infection: no recorded evidence of infection prior to MSD sample or within 1 week after, and MSD for anti-nucleocapsid antibodies <1200.

2. Pre-vaccination infection: Positive test for active infection or presence of anti-nucleocapsid antibodies, or admission to hospital with confirmed or suspected COVID-19, on or before date of second vaccine dose.

3. Post-vaccination breakthrough infection: Positive test for active infection, or admission to hospital with confirmed or suspected COVID-19, after date of second vaccine dose, with no evidence of infection having occurred prior to second vaccine dose.

4. Abbott antibodies detected: Antibodies detected using Abbott nucleocapsid assay prior to MSD sample or within 1 week after (to allow for small variations in sample and test dates), with no evidence of infection prior to second vaccine dose and without direct evidence of active infection after second vaccine dose. These cases could reflect either infection prior to vaccination or breakthrough infection that was not detected and/or recorded.

5. MSD nucleocapsid-antibodies detected: No recorded evidence of infection prior to MSD sample or within 1 week after, but MSD for anti-nucleocapsid antibodies ≥1200 for index or prior sample. These cases are classified as having experienced a past SARS-CoV-2 infection on the basis of the MSD anti-nucleocapsid assay.

Individuals from the main ‘linked’ dataset could be included within any of these categories. Those not in the ‘linked’ dataset lacked a linked record of testing history for SARS-CoV-2 or of hospital admissions but did have a record of serial blood samples subject to MSD and Abbott anti-nucleocapsid antibody assay testing. They could therefore be included in any group except (3), although inclusion in group (2) could only be on the basis of Abbott anti-nucleocapsid antibody assay testing where available.

#### **Table S1:** Breakdown of individuals by vaccine type, sex, and infection classification at first MSD observation by resident/staff status

|  | ***Resident,* n (%)** | ***Staff,* n(%)** | ***Total,* n(%)** |
| --- | --- | --- | --- |
| *Vaccine* |  |  |  |
| Oxford-AstraZeneca | 254 (63.2) | 242 (38.3) | 496 (48.0) |
| Pfizer-BioNTech | 148 (36.8) | 390 (61.7) | 538 (52.0) |
| *Sex* |  |  |  |
| Female | 282 (70.2) | 550 (87.0) | 832 (80.5) |
| Male | 120 (29.9) | 82 (13.0) | 202 (19.5) |
| *Infection category at first observation* |  |  |  |
| 1. No evidence of infection | 164 (40.8) | 311 (49.2) | 475 (45.9) |
| 2. Infection before vaccination | 123 (30.6) | 69 (10.9) | 192 (18.6) |
| 3. Breakthrough infection | 4 (1.0) | 3 (0.5) | 7 (0.7) |
| 4. Antibodies on Abbott anti-nucleocapsid assay, without evidence of active infection | 44 (11.0) | 73 (11.6) | 117 (11.3) |
| 5. Antibodies on MSD anti-nucleocapsid assay, without other evidence | 67 (16.7) | 176 (27.9) | 243 (23.5) |
| *Data source* |  |  |  |
| Full linkage to Vivaldi dataset | 333 (82.8) | 262 (41.5) | 595 (57.5) |
| Additional data without test history | 69 (17.2) | 370 (58.5) | 439 (42.5) |

A total of 243/718 (33.8%) individuals with no other evidence of previous SAR-CoV-2 infection have been reclassified as having had a prior infection on the basis of their initial MSD anti-nucleocapsid antibody result. Assuming a specificity of 96% (and allowing a uniform prior for the true seroprevalence in this group), we would expect 20 of the reclassified individuals to represent false-positives (95% prediction interval 10-29).

#### **Table S2:** Estimated marginal intercept, slope terms and corresponding half-life from the final statistical model for anti-spike antibody levels for participant sub-groups

| **Subject** | **Vaccine** | **Sex and inf. status** | ***Average peak (intercept) (95%CI)*** | ***Average value at 6 months*(95%CI)*** | ***Slope (95%CI) (annual)*** | ***Half-life (days)*** |
| --- | --- | --- | --- | --- | --- | --- |
| *Resident* | *Ox.-AZ* | *Female: no inf.* | 4.07 (3.84 to 4.29) | 3.75 (3.56 to 3.95) | -0.63 (-1.33 to 0.06) | 174 (83 to inf) |
|  |  | *Female: prior inf.* | 4.75 (4.55 to 4.95) | 4.68 (4.51 to 4.85) | -0.14 (-0.73 to 0.46) | 806 (151 to inf) |
|  |  | *Male: no inf* | 4.30 (4.07 to 4.53) | 3.58 (3.39 to 3.77) | -1.45 (-2.12 to -0.78) | 76 (52 to 140) |
|  |  | *Male: prior inf.* | 4.94 (4.7 to 5.19) | 4.50 (4.29 to 4.72) | -0.88 (-1.6 to -0.15) | 125 (68 to 741) |
|  | *Pfizer-B* | *Female: no inf.* | 4.99 (4.73 to 5.26) | 4.07 (3.88 to 4.25) | -1.85 (-2.48 to -1.22) | 59 (44 to 90) |
|  |  | *Female: prior inf.* | 5.42 (5.22 to 5.62) | 4.72 (4.57 to 4.87) | -1.40 (-1.9 to -0.91) | 78 (58 to 121) |
|  |  | *Male: no inf* | 5.08 (4.82 to 5.35) | 4.05 (3.87 to 4.24) | -2.06 (-2.68 to -1.44) | 53 (41 to 76) |
|  |  | *Male: prior inf.* | 5.51 (5.29 to 5.74) | 4.71 (4.55 to 4.86) | -1.62 (-2.16 to -1.07) | 68 (51 to 102) |
| *Staff* | *Ox.-AZ* | *Female: no inf.* | 4.33 (4.16 to 4.49) | 3.83 (3.69 to 3.97) | -1.00 (-1.5 to -0.5) | 110 (73 to 221) |
|  |  | *Female: prior inf.* | 5.00 (4.86 to 5.15) | 4.75 (4.62 to 4.89) | -0.50 (-0.93 to -0.07) | 219 (118 to 1562) |
|  |  | *Male: no inf* | 4.50 (4.25 to 4.74) | 3.65 (3.44 to 3.86) | -1.69 (-2.41 to -0.96) | 65 (46 to 115) |
|  |  | *Male: prior inf.* | 5.17 (4.93 to 5.42) | 4.58 (4.37 to 4.79) | -1.19 (-1.91 to -0.46) | 92 (57 to 237) |
|  | *Pfizer-B* | *Female: no inf.* | 5.04 (4.9 to 5.17) | 4.22 (4.12 to 4.31) | -1.64 (-1.98 to -1.3) | 67 (55 to 84) |
|  |  | *Female: prior inf.* | 5.48 (5.34 to 5.61) | 4.87 (4.78 to 4.97) | -1.21 (-1.56 to -0.87) | 91 (71 to 127) |
|  |  | *Male: no inf* | 5.14 (4.93 to 5.36) | 4.21 (4.05 to 4.36) | -1.87 (-2.37 to -1.37) | 59 (46 to 80) |
|  |  | *Male: prior inf.* | 5.58 (5.35 to 5.82) | 4.86 (4.7 to 5.03) | -1.44 (-2.02 to -0.86) | 76 (54 to 127) |

Staff age set at 50 and resident age at 86

*From peak level 21 days after second vaccine dose.

**Figure S1:** Log10-transformed MSD values for anti-spike antibody levels in relation to the time from second vaccine dose, divided by vaccine type and staff/resident status, and colour-coded by prior infection category (observations from the same person linked by lines)

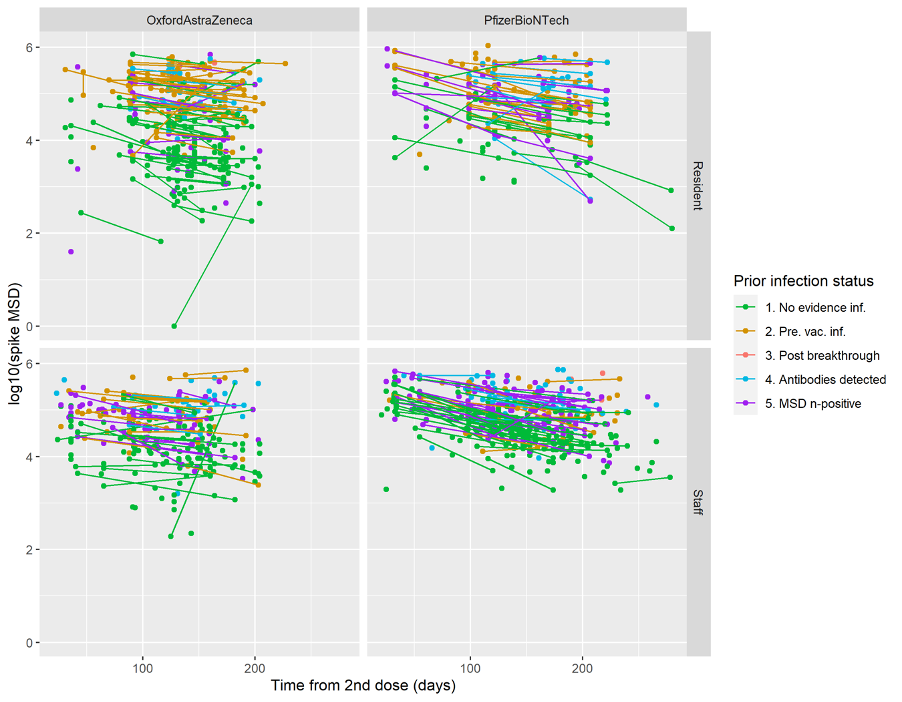

**Figure S2:** MSD values for anti-spike antibody levels in relation to the time from second vaccine dose, divided by vaccine type and staff/resident status, and colour-coded by simplified prior infection category as used in the statistical analysis (observations from the same person linked by lines)

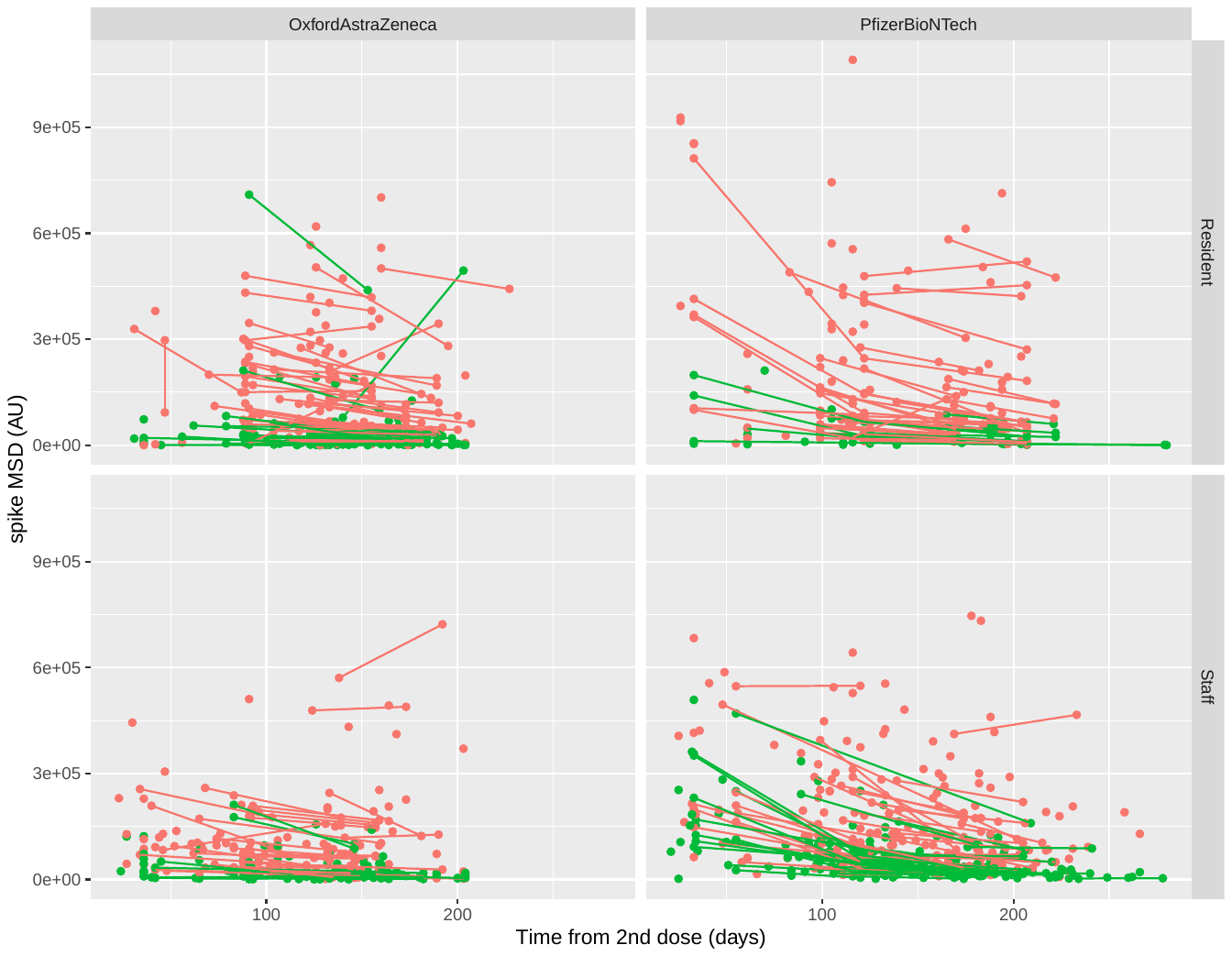

Observations with evidence of prior infection are shown in red, whilst those without are shown in green.

### **MSD data for anti-spike antibody level in relation to anti-nucleocapsid level**

**Figure S3:** Pairwise associations between MSD antibody levels for spike against those for nucleocapsid for the first post-vaccination sample included for each of the 1034 individuals
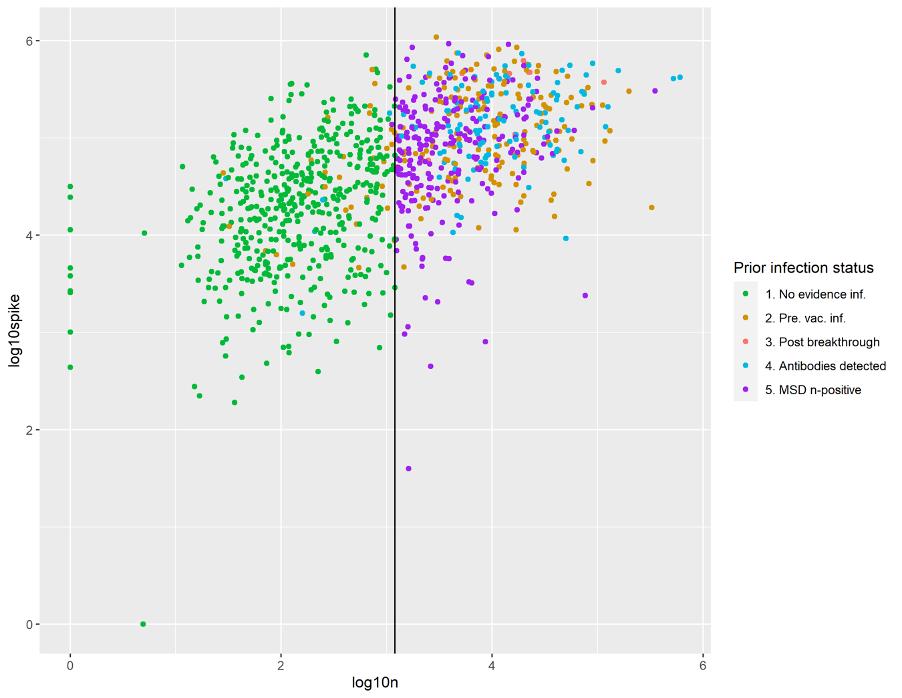
The vertical line shows the cut-off of 1200 AU/mL for nucleocapsid (n).

A large majority (470/505, 93.1%) of MSD observations with anti-nucleocapsid antibody response <1200 were from individuals with no evidence of prior SARS-CoV-2 infection at point of sampling. However, 242/712 (34.0%) of those with no evidence of prior SARS-CoV-2 infection from other sources of information had a anti-nucleocapsid antibody response ≥1200, with clear overlap in the distribution with those individuals who did have evidence of prior infection. Seven individuals were missing an MSD value for anti-nucleocapsid antibody level at first observation. Those with high anti-nucleocapsid MSD levels also mostly have high anti-spike MSD levels, indicating the boosting of anti-spike antibody levels through undetected (or unlinked evidence of) infection in a substantial proportion of these patients.

### **MSD data in comparison to Abbott results**

#### **Figure S4:** Quantitative Abbott anti-nucleocapsid antibody titres vs MSD anti-nucleocapsid antibody titres from the first post-vaccination sample for 380 residents and 578 staff

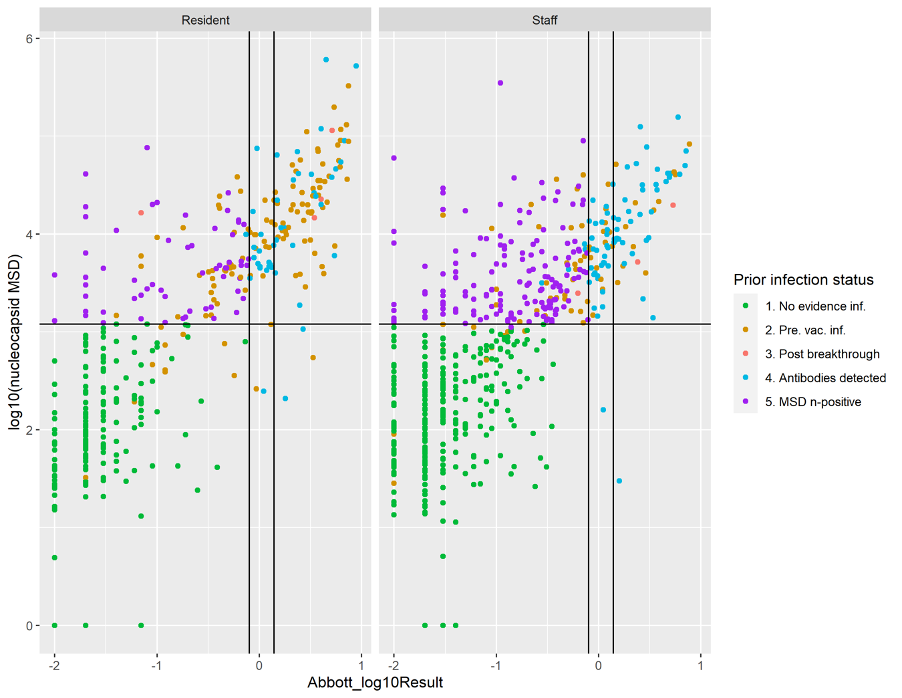

The threshold of 1200 is shown for the MSD results (horizontal line), with thresholds of 0.8 (cut-off used for our research) and 1.4 (manufacturer’s cut-off) shown for the Abbott results (vertical lines), with color-coding by ‘infection category’.

There is a clear positive correlation between Abbott scores and the MSD assay results for anti-nucleocapsid antibody levels, with substantial numbers of individuals with evidence of prior infection falling below either threshold for the Abbott test.

### **Anti-nucleocapsid antibodies in relation to time from infection**

**Figure S5:** MSD anti-nucleocapsid antibody titres vs time from most recent known active infection (horizontal line=log10(1200)) [107 observations from 78 residents and 47 from 34 staff]

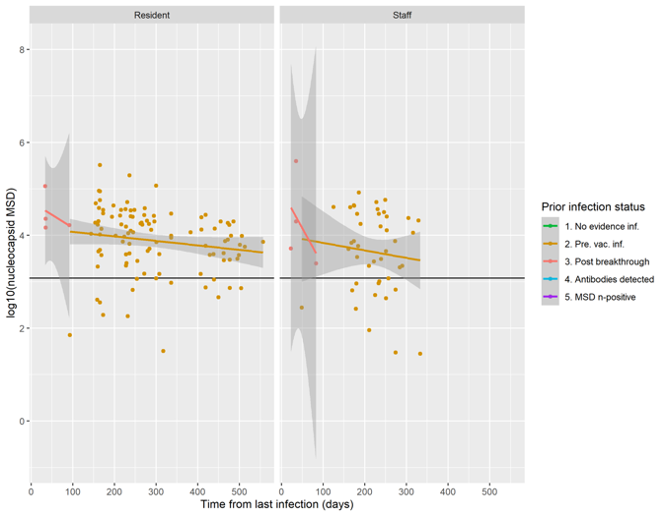

**Figure S6:** Abbott anti-nucleocapsid antibody titres vs time since most recent known active infection (horizontal lines=log10(0.8) and log10(1.4)) [104 observations from 78 residents and 45 from 34 staff]

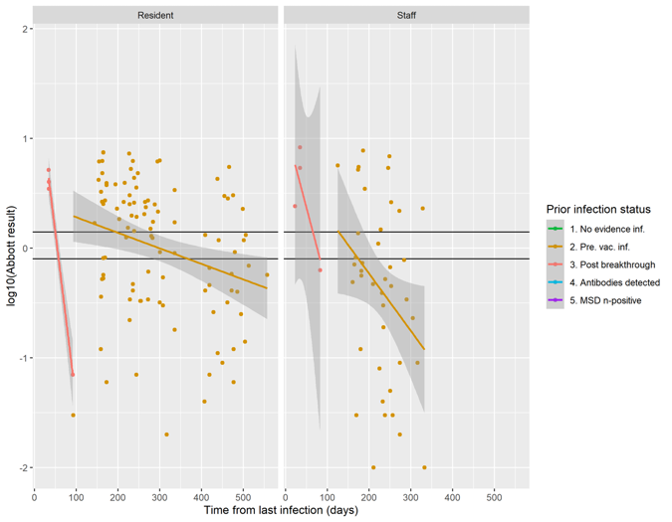

The sensitivity of the Abbott test drops substantially with time from infection, whereas the drop in sensitivity using the MSD anti-nucleocapsid antibody assay is relatively small up to 18 months from time of infection.

### **STROBE Checklist**

|  | **Item No** | **Recommendation** | **Page No** |
| --- | --- | --- | --- |
| Title and abstract | | |  |
|  | 1 | (*a*) Indicate the study's design with a commonly used term in the title or the abstract | 2 |
|  |  | (*b*) Provide in the abstract an informative and balanced summary of what was done and what was found | 2 |
| Introduction | | |  |
| Background/rationale | 2 | Explain the scientific background and rationale for the investigation being reported | 2, 3 |
| Objectives | 3 | State specific objectives, including any prespecified hypotheses | 3 |
| Methods | | |  |
| Study design | 4 | Present key elements of study design early in the paper | 3 |
| Setting | 5 | Describe the setting, locations, and relevant dates, including periods of recruitment, exposure, follow-up, and data collection | 3 |
| Participants | 6 | *a) Cohort study*? Give the eligibility criteria, and the sources and methods of selection of participants. Describe methods of follow-up  *Case-control study*? Give the eligibility criteria, and the sources and methods of case ascertainment and control selection. Give the rationale for the choice of cases and controls  *Cross sectional study*? Give the eligibility criteria, and the sources and methods of selection of participants | 3 |
|  |  | (*b*) *Cohort study*? For matched studies, give matching criteria and number of exposed and unexposed  *Case-control study*? For matched studies, give matching criteria and the number of controls per case |  |
| Variables | 7 | Clearly define all outcomes, exposures, predictors, potential confounders, and effect modifiers. Give diagnostic criteria, if applicable | 3, 4 |
| Data sources/ measurement | 8* | For each variable of interest, give sources of data and details of methods of assessment (measurement). Describe comparability of assessment methods if there is more than one group | 3 |
| Bias | 9 | Describe any efforts to address potential sources of bias | 3, 5 |
| Study size | 10 | Explain how the study size was arrived at | 4 |
| Quantitative variables | 11 | Explain how quantitative variables were handled in the analyses. If applicable, describe which groupings were chosen and why | 3, 4 |
| Statistical methods | 12 | (*a*) Describe all statistical methods, including those used to control for confounding | 3, 4 |
|  |  | (*b*) Describe any methods used to examine subgroups and interactions | 3, 4 |
|  |  | (*c*) Explain how missing data were addressed | 3, 4 |
|  |  | (*d*) *Cohort study*? If applicable, explain how loss to follow-up was addressed  *Case-control study*? If applicable, explain how matching of cases and controls was addressed  *Cross sectional study*? If applicable, describe analytical methods taking account of sampling strategy |  |
|  |  | (*e*) Describe any sensitivity analyses | n/a |
| Results | | |  |
| Participants | 13* | (*a*) Report numbers of individuals at each stage of study? eg numbers potentially eligible, examined for eligibility, confirmed eligible, included in the study, completing follow-up, and analysed | 4 |
|  |  | (*b*) Give reasons for non-participation at each stage | 4, S1 |
|  |  | (*c*) Consider use of a flow diagram | S1 |
| Descriptive data | 14* | (*a*)Give characteristics of study participants (eg demographic, clinical, social) and information on exposures and potential confounders | 4, S1 |
|  |  | (*b*) Indicate number of participants with missing data for each variable of interest | S1 |
|  |  | (*c*) *Cohort study*? Summarise follow-up time (eg average and total amount) | 4 |
| Outcome data | 15* | *Cohort study*? Report numbers of outcome events or summary measures over time | 4, tables |
|  |  | *Case-control study?* Report numbers in each exposure category, or summary measures of exposure |  |
|  |  | *Cross sectional study?* Report numbers of outcome events or summary measures |  |
| Main results | 16 | (*a*) Report the numbers of individuals at each stage of the study?eg numbers potentially eligible, examined for eligibility, confirmed eligible, included in the study, completing follow-up, and analysed | 4 |
|  |  | (*b*) Give reasons for non-participation at each stage | 4, S1 |
|  |  | (*c*) Consider use of a flow diagram |  |
| Other analyses | 17 | Report other analyses done?eg analyses of subgroups and interactions, and sensitivity analyses | n/a |
| Discussion | | |  |
| Key results | 18 | Summarise key results with reference to study objectives | 4, 5 |
| Limitations | 19 | Discuss limitations of the study, taking into account sources of potential bias or imprecision. Discuss both direction and magnitude of any potential bias | 5 |
| Interpretation | 20 | Give a cautious overall interpretation of results considering objectives, limitations, multiplicity of analyses, results from similar studies, and other relevant evidence | 5 |
| Generalisability | 21 | Discuss the generalisability (external validity) of the study results | 5 |
| Other information | | |  |
| Funding | 22 | Give the source of funding and the role of the funders for the present study and, if applicable, for the original study on which the present article is based | 1 |
